# Metaphors that shape parents’ perceptions of effective communication with healthcare practitioners following child death: a qualitative UK study

**DOI:** 10.1101/2021.07.08.21259704

**Authors:** Sarah Turner, Jeannette Littlemore, Annie Topping, Eloise Parr, Julie Taylor

**Affiliations:** School of Humanities, Coventry University, Coventry, UK; Department of English Language and Linguistics, University of Birmingham, Birmingham, UK; School of Nursing, University of Birmingham, Birmingham, UK; University Hospitals Birmingham NHS Foundation Trust, Birmingham, UK; Birmingham Women’s and Children’s Hospitals NHS Trust, Birmingham, UK

**Author notes:** Corresponding author address: College of Medical and Dental Science, University of Birmingham, Edgbaston, B15 2TT, UK. Lead author address: School of Humanities, Coventry University, Coventry, UK.

**Keywords:** bereavement, communication, metaphor, child death, healthcare professionals, parents

## Abstract

**Objectives:** To offer an interpretation of bereaved parents’ evaluations of communication with healthcare practitioners surrounding the death of a child.

**Design:** Interpretative qualitative study employing thematic and linguistic analyses of metaphor embedded in interview data.

**Setting:** England and Scotland.

**Participants:** 24 bereaved parents (21 women, 3 men)

**Methods:** Participants were recruited through the True Colours Trust website and mailing list, similar UK charities, and word of mouth. Following interviews in person or via video-conferencing platforms (Skype/Zoom), transcripts first underwent thematic and subsequently linguistic analyses supported by Nvivo. A focused analysis of metaphors used by the parents was undertaken allow in-depth interpretation of how they conceptualised their experiences.

**Results:** The findings illuminate the ways parents experienced communication with healthcare practitioners surrounding the death of a child. They show how their evaluations of effective care relate to the experience of the bereavement itself, as expressed through metaphor. We identified three broad themes: (1) Identity (2) Emotional and Physical Response(s) and (3) Time. Successful communication from healthcare practitioners takes account of parents’ experiences related to these themes.

**Conclusions:** This study suggests that healthcare practitioners when communicating with bereaved parents need to recognise, and seek to comprehend, the ways in which the loss impacts upon an individual’s identity as a parent, the ‘physical’ nature of the emotions that can be unleashed, and the ways in which the death of a child can alter their metaphorical conceptions of time.

**Strengths and limitations of the study:**

- This interpretative qualitative study contributes to the growing literature on the experience of parental bereavement through its focus on the use of metaphor in parents’ accounts of child loss.
- The metaphor analysis afforded a focus not only on the content of the interviews, but also on the language that was used to express this content, providing more depth and nuance to the interpretation of the findings.
- Participants were all UK-based and self-selecting which could limit transferability.

## INTRODUCTION

When parents experience the death of a child, no matter the age, they experience a range of complex, often intolerably painful emotions[1]. The experience and effects of the death of a child on parents has received considerable attention, including the psychosocial impacts experienced as a consequence of devastating and enduring grief[2]; the experience of parents around end of life of a child[3–5]; what they want and expect from healthcare practitioners (HCPs)[2,6]; and impacts of bereavement services and interventions[1,7]. Less work has focused on the communication remembered by parents as part of their experiences of care or services received, or how that communication subsequently scaffolds their experience of bereavement. However, negative interactions with HCPs following the death of a child have been shown to exacerbate the trauma experienced by the parents[8], while positive interactions such as empathy and provision of information can mitigate it[9,10]. A better understanding of the impact of bereavement and the ways in which it affects parents’ evaluations of the communication they had with HCPs in the period surrounding the death may enable design of structures, processes, interventions and training to improve individualised compassionate bereavement care.

## BACKGROUND

The death of a child engenders a range of intense and complex emotions that may be difficult to articulate[11]. The loss of a child has been associated with poorer parental physical health and increased mortality[12]. Staff regularly providing end-of life care also experience high levels of emotional exhaustion[5]. These are further compounded when HCPs feel guilt about inadequate, or missed care, poor communication and/or contain expressions of grief or emotions in order to protect families from further distress[13]. Effective communication following the death of a child is therefore important for both parents and HCPs.

Staff involved in managing deaths occurring in healthcare settings have well practiced roles and responsibilities. Sociocultural values and beliefs about death and dying are implicitly embedded in the organisation of services and inevitably demonstrated in the behaviours, attitudes and discourses used by the staff involved[14]. This is most evident in what could be interpreted as a proceduralisation of dying[15,16] which can exclude the individual who is dying and parents and family members experiencing the loss. The well rehearsed concept of a good death is less well understood in relation to the death of a child or young person[17]. Possibly as greater emphasis is placed on dying and/or bereavement rather than the period surrounding death, and different participants (patients, family/parents, and staff) may have different needs and requirements for control and participation/non-participation in the social context where the dying and death are enacted.

Parents provide mixed accounts of the care that they recall having received in the period surrounding the death of their child[2,18] with some reporting highly positive experiences of compassionate care and others less positive accounts. What is clear is parents’ value open and honest communication with HCPs, want to be actively involved, and their emotional distress acknowledged. Ideally they want to communicate with HCPs with whom they have already developed established and trusting relationships[4]. Furthermore, they want to say goodbye, to understand why their child died and to receive follow up care from HCPs involved in the care of their child[2]. Ensuring these standards are achieved may be particularly challenging in the case of sudden death or in emergency, trauma and critical care contexts. This means that good guidance for healthcare practitioners on how best to communicate with parents following the death of their child is essential.

In this study, we focused on the experiences of parents in the immediate aftermath of the death of their child, and their recollected communications with HCPs. We included parents of both adult and non-adult children. The term ‘healthcare practitioners’ (HCPs) is used in a broad sense to include all of those who come into contact with the parents at the hospital, not just the doctors and nurses.

## METHODS

### Aim

The aim of this research was to investigate how parents’ experiences of bereavement and child death shaped their perceptions of the communication they had with HCPs. In this paper, we consider ‘communication’ to encompass both language and behaviour, and the interactions between actors.

By gaining insights into parents’ experiences of child death, we sought to better understand what constitutes effective communication, with a view to improving care after loss.

### Design and Setting

We used an interpretative descriptive qualitative design[19]. The study is reported using Consolidated criteria for Reporting Qualitative research guidelines (see online supplementary file).

### Participants

Our participants comprised parents aged at least 18 years who had lost a child of any age, who could speak English at conversational level, and who were able to provide informed consent.

### Sampling and recruitment

A purposive sampling procedure was employed. Recruitment adverts were posted by the True Colours Trust, Compassionate Friends, and other similar charities (e.g. Acorns Children’s Hospice, Winston’s Wish etc). Potential participants who responded to the posting were contacted by the research team and invited to participate in an interview.

### Data collection

In total, 24 participants were interviewed, in 21 interviews. The majority of the interviews were conducted on a 1-1 basis, with all authors undertaking interviews. On three occasions however we undertook joint interviews with male and female participants at their request (interviews 2, 5 and 20). All interviews involved female participants (n=21). Interestingly just under half of our respondents were themselves HCPs. The interviews were conducted face to face (n=8), or via video-conference platforms (n= 13), according to participant preference, location, or COVID-19 restrictions. All but two of the face to face interviews were conducted in participants’ homes, except two conducted at the Charity headquarters. Interviews lasted an average of 81 minutes (range: 34-129 mins).

Prior to interview, participants were informed how the findings derived from their interview data would be reported and disseminated in training materials. All participants provided informed consent prior to interview. Interviews were undertaken by all five authors, with some interviewers working together. A topic guide was developed to serve as a framework for the semi-structured interviews (see box 1). Open questions were used to encourage extended answers, and to allow the interviewer(s) to elicit further detail on points of interest.

#### Box 1

**Sample of interview prompts**

- Can you tell us about [name of child]?
- Please could you describe what happened ….?
- Please could you tell me about what happened next…?
- How did you feel about…?
- Can you tell me about the communication you had with the healthcare practitioners, funeral directors and registrars?
- Can you recall any examples of particularly sensitive and effective communication you received?
- Can you recall any examples of particularly insensitive communication you received?
- Imagine you are talking to someone who supports people who have had an experience like yours. What advice would you give them?
- What would you like to see changed?

Audio recordings were made of the interviews with consent. In the case of video-conference interviews, recordings included video data, but these were destroyed and only audio recordings retained.

### Analysis

Audio recordings were transcribed verbatim and all identifying information removed. Transcripts were sent to the participants to allow them to make any corrections or additions they felt necessary. Interview transcripts were coded using NVivo.

We first undertook a thematic analysis to identify parents’ experiences of bereavement and communication. A full list of identified themes is provided in the online supplementary file 1]. Subsequently, we combined our thematic analysis with a metaphor analysis which enabled us to explore how parents conceptualised these experiences. The metaphor analysis involved coding the language used by participants relating to each of the categories for metaphor. This focus on metaphors was designed to provide deeper insights into the nature of the experience, and explore the extent to which parents’ perceptions of communication could be explained by these experiences.

In order to identify metaphor in the transcripts, we used Cameron and Maslen’s Vehicle Identification Procedure[20]. This analytic procedure operates at the level of the phrase and involves recognition of the fact that something is being described in terms of a different entity.

### Patient and Public Involvement

The study engaged with bereaved families throughout. It was designed in communication with the True Colours Trust who funded the research, and who work closely with bereaved parents. Study results will be shared with all participants and ultimately integrated in training materials.

## FINDINGS

### Participants

Twenty-four people participated in 21 interviews. Table 1 gives further information about the circumstances of death in each interview.

**Table 1:** Circumstances of death

| Interview number | Age of child (years) | Cause of death | When death occurred |
| --- | --- | --- | --- |
| 1 | 0-5 | RTC | <5yrs |
| 2 | 0-5 | Illness (sudden) | >5yrs |
| 3 | 16-20 | Illness (sudden) | >5yrs |
| 4 | 0-5 | Illness (sudden) | >5yrs |
| 5 | 16-20 | Drug-related death | <5yrs |
| 6 | 0-5 | Illness (sudden) | <5yrs |
| 7* | 0-5 | Illness (genetic condition) | >5yrs + <5yrs |
| 8 | 0-5 | Illness (sudden) | >5yrs |
| 9 | 16-20 | Suicide | >5yrs |
| 10 | 21-30 | Suicide | >5yrs |
| 11 | 16-20 | Drug-related death | <5yrs |
| 12 | 21-30 | Unknown (ongoing inquiry) | <5yrs |
| 13 | 21-30 | Suicide | <5yrs |
| 14 | 21-30 | Illness (cancer) | <5yrs |
| 15 | 16-20 | RTC | >5yrs |
| 16 | 21-30 | Murder | <5yrs |
| 17 | 0-5 | Illness (sudden) | <5yrs |
| 18 | 0-5 | Illness (sudden) | >5yrs |
| 19 | 0-5 | Illness (genetic condition) | >5yrs |
| 20 | 0-5 | Illness (sudden) | <5yrs |
| 21 | 0-5 | Illness (sudden) | >5yrs |

\* interviewee discusses two child deaths associated with the same inherited condition

In all our interviews, we found that the parents had vivid memories of some parts of the peri-death experience whereas others were more hazy. In all cases, the ways in which HCPs communicated with the parents during this period played a crucial role in shaping the experience. In this example, the parent uses metaphor to illustrate the painful impact of the doctor’s words:

> years later, I can remember every word. I can even remember his tone of voice as he said it. Absolutely etched with acid into my brain
>
> [Interview 3]

Through our thematic and metaphorical analyses we identified 14 categories, which included different kinds of communication, people involved, and experiences surrounding events associated with the loss (e.g. funerals). These are shown in Table 1 in the supplementary file.

To answer our research questions we examined the metaphors within these categories in order to identify overarching themes in the data where metaphor was playing an important role. The three themes reported here and shown in Figure 1 are: (1) Identity (2) Emotional and Physical Response and (3) Time.

**Figure 1:**
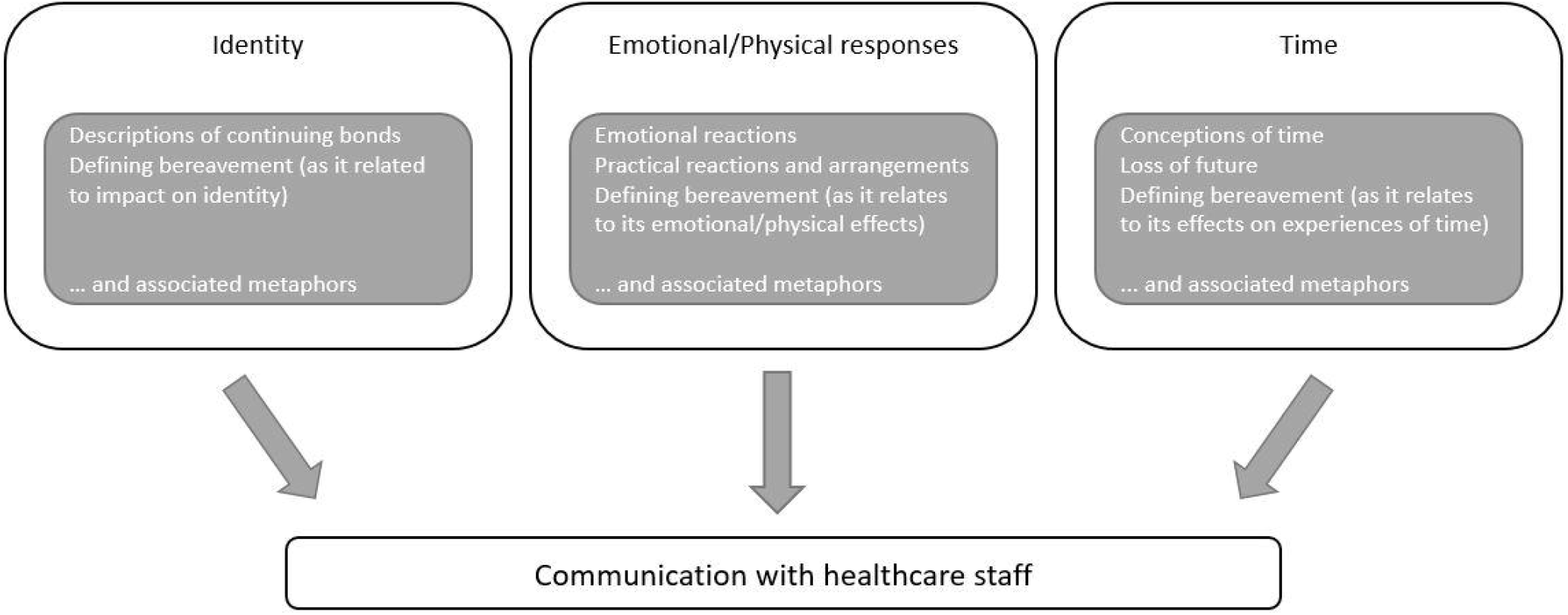
Metaphorical Themes

### Identities of parents and children

Parents appeared to struggle with the loss not only as the death of the child but also as a significant threat to their identities as parents. Some parents talked about how leaving their child in the hospital after death was made more difficult, as it constituted ‘leaving them behind’. Parents felt that they still needed to look after their child, as one parent explained how she was ‘distraught’ that her daughter was ‘shut up’ in the mortuary as she was claustrophobic. These utterances reflect a level of metaphoric thinking in which parents conceptualise their child as being, on some level, still alive. Here a bereaved parent talked about the excellent care received when they went to identify their son’s body:

> [a member of staff at the mortuary] crouched down, she took my hand and said ‘hello NAME, I’ve got your little boy and I’m keeping him safe, do you want to come and see him?’ [Interview 5]

Many of the parents commented on how they were given the time and space to act in ways that seemed right to them as it respected their child’s identity and theirs as parents. One couple (Interview 2) appreciated how the consultant arranged for their child’s body to be taken to a children’s hospice, where it could be kept in a refrigerated room that had been decorated like a child’s bedroom.

Conversely, some parents reported examples of care and communication that failed to recognise their child as an individual. This parent describes the distress she felt when hospital staff attempted to fit her young toddler into a Moses basket:

> They tried to put him in a Moses basket cos they thought it was a cot death, they obviously didn’t say the age on the phone call across […] They probably should know about the age of the child, that was really – cos they did try and like squeeze him in in a way as well. So even when – you could clearly see he wasn’t gonna fit in that. I just thought that was quite upsetting. Like he wouldn’t have been in a Moses basket at that age anyway [Interview 2].

This was particularly distressing to the parents because it seemed that the HCPs were more concerned with following the procedure rather than with responding flexibly to the reality of the situation. This contributed to the parents’ impression that the care their child received was a “one size fits all” rather than tailored to the individual needs of their child.

In some cases, parents were told that they were not allowed to interact with their child following the death because the circumstances of the death were unknown. Although necessary, it caused extreme distress, exacerbated by insufficient explanations and/or management of parental expectations. Another couple (Interview 6) spoke of the ‘horrific’ experience of having to take a service lift down to the ‘old-fashioned’ and ‘cold’ mortuary. The parents felt that they were insufficiently prepared for what they would encounter, and their role as parents was discounted to that of observers of a body.

### Emotional and Physical Responses

A second theme related to the ‘physical’ nature of emotions unleashed by the death of a child. Parents often referred to these experiences in metaphorical ways conveying the strength of their emotional response to the death.

> You’re frozen, you’re like a zombie, you just can’t, everything’s frozen up inside you
>
> [Interview 3]
>
> It’s like you’re full of cotton wool in your head and people do have to keep telling you things over and over and over again … so just kind of time, don’t be surprised you’ve got a woolen brain sitting in front of you
>
> [Interview 3]

These quotes emphasise the effect death has and how HCPs may need to recognise this and in terms of ability of parents to receive information. In the following example, a parent talks about how she had kept a bereavement support leaflet from the hospital for some months following her daughter’s death.

> It was there in the booklet that HOSPITAL had given me […] my attention was directed to it and I hadn’t noticed it. I’d been to my doctor and he’d given me sleeping tablets and I wasn’t taking them, I was saving them up. It was stupid, I know it was stupid. [I was] sitting with all these tablets and a bottle of wine and just thinking, ‘I can’t do this’. … And I thought I’m gonna have to get help and I knew the Samaritans number was in that booklet they’d given me. So I’d found the booklet, found the Samaritans and then just underneath I saw Compassionate Friends. And so I phoned the helpline … and got an answerphone, ‘We’ll get back to you’ and I thought, ‘I bet they don’t’, but they did. And I spoke to a lady called NAME … and she was great. She’d lost her daughter about 14 or 15 years before and just hearing her say that she’d survived more than anything else helped me.
>
> [Interview 3]

At times parents employed metaphor to express the fact that they no longer wanted to engage with the world:

> We just wanted to hide. Literally had this sense of ‘I wanna get a cave, I don’t wanna see the world, I don’t wanna engage with anybody or anything’. I can’t function with the world being normal. I nearly always wore contact lenses, I rarely wore glasses but [then] I wore glasses on purpose so that I could just take them off half the time, cos I just wanted everything to go away [Interview 4].

She draws a metaphorical comparison between the act of seeing and the act of engaging with the world around her, which allows her to take control of her situation by literally removing her glasses, thus enabling her to metaphorically withdraw from the world.

At times, parents commented that HCPs appeared not to take sufficient account of the fact that appearing calm concealed the depth of their distress, how staff mistook calmness for strength and indeed congratulated parents for coping. Whereas the bereaved felt under pressure to present a ‘brave face’:

> Throughout the early days people (hospital staff etc.,) would constantly say how strong and brave I was. I kept saying “no I’m not, I’m just pretending”, but eventually I felt that I had to be this “brave, strong” woman or I would be letting everyone down. If I could say one thing to them [hospital staff] it would be to ask them not to say that. We are not brave and strong, just in a state of severe shock and watching ourselves go through the motions and it’s really unhelpful to make us feel we must behave in a certain way [Interview 11].

By praising parents HCPs were inadvertently reinforcing the idea that following bereavement, some kinds of behaviour tend to be more acceptable than others and that overt emotional displays are to be discouraged[4].

### Time

Previous studies have shown that trauma can exert a strong effect on perceptions of time, reporting the phenomena of time stopping, slowing down, speeding up or becoming irrelevant[21]. All of these descriptions draw on the metaphorical relationship between time and movement through space[22]. Time appeared to be experienced differently by parents on the death of a child, and time becomes foregrounded in their experience. This is demonstrated in the following examples:

> So suddenly … I remember thinking, ‘Slow down! It’s like I still don’t know what’s happened,’ and then suddenly, we were like, ‘Whoo!’ and we were heading towards an inquest but having said that, it was a year, this whole process … so it wasn’t slow down time. It was just - so we had no control [Interview 1].
>
> when you’ve had such a massive shock like that, it just takes so long for your brain to catch up [Interview 1].

Disordered time appeared to influence how interactions with HCPs were perceived. For example what could be viewed as acts of caring for were interpreted as simply procedural and are therefore less effective. In some cases, this may prevent parents from reaping any benefit from the care provided. For example, one participant described receiving a memory box without any (remembered) explanation of what it was, or what was in it, and did not dare to open it:

> At that point, a nurse put something in my mum’s hands and it was inside a cardboard box and just said, ‘Here’s his memory box’. So my mum was a bit like, ‘Thanks’. It was horrific for her. So she walked out, there was no other explanation of it and we walked out and she just put it on the dining table when we got home. And like for days after everyone was like, ‘What’s in that box?’, but no one dared open it because - my mum was saying, ‘They said oh it’s his memory box, I didn’t want to open it in case it’s his things in there’ [Interview 6].

We see from this example that what may have started out as an enactment of good practice in bereavement support - the giving of a memory box - had become for this parent, something to be feared due to the way in which it was presented.

## DISCUSSION

The findings from this study suggest that good communication with HCPs following the death of a child should acknowledge parental identity (and that of their child as an individual) and offer opportunities for them to enact this; taking account their emotional and physical experiences; and accommodate their altered experiences of time. Practitioners need to acknowledge the individual needs of the parents while working within the parameters of best practice and legal considerations[16,23].

### Identity

In keeping with previous research[24,25], we found that many of the parents reported a strong and sustained bond with their child, which led to a continuing need to ‘parent’ their child. The importance of these bonds may be understood as a response to the threat to parental identity that the loss of a child represents. Thus parents experiencing their child’s death are in part losing their role as a parent [26]. Parents in our study highlighted the importance of being able to spend time with their child following death and to continue enacting their roles as parents[27–29]. The opportunity to do this allows for a ‘slow and gentle separation’ at a pace set by the parents, reduced feelings of helplessness, and the potential to form ‘bittersweet memories’ which may later represent a source of comfort[27,28]. The importance of protected time within the grief process particularly spending time with the child’s body represents symbolically privileged status of parent and other family members in death[30].

The behaviour of parents towards their child’s body can be understood as an example of metaphorical thinking, where the child is perceived as needing the same care as in life. This involves a blurring of the boundaries between life and death where parents enact behaviours they would have engaged in if the child were alive. Our study provides further evidence of the importance of opportunities to engage in these behaviours, and their facilitative role in the grieving process.

HCPs have an important role to play in acknowledging bereaved parents’ continuing bonds with their children, and facilitating opportunities for parents to perform these behaviours. The parents in our study made frequent references to cases where HCPs were either successful in doing so, or where they felt the care they received was lacking in this regard. This was often a determining factor in their evaluation of the effectiveness of the overall care that they had received. For example, parents in our study appreciated instances of HCPs treating their child’s body with respect and dignity. This was represented in our findings describing HCPs speaking to children after death as a means of respecting the child’s presence and personhood[31]. Effective linguistic and behavioural communication therefore involves recognising that the loss of a child represents a significant threat to parental identity, and that parents should be given the time, space and opportunity to engage in activities that enable them to enact that identity. Similarly, there is a need to understand the continuing bonds that the parents still have with their child, even when these may be appear in ways that seem unusual.

### Emotional and physical response

The parents in our study drew heavily on embodied metaphors to describe the strength and impact of the bereavement and ensuing emotions. The metaphors used had such a strong physical basis is in line with other studies that propose powerful, embodied metaphors can be the only device strong enough to to express emotions following traumatic experiences[32,33,33–36]. Although metaphor is sometimes seen as a poetic device, operating solely at the level of language, we have demonstrated that it is inappropriate to see it as a surface-level linguistic phenomenon, particularly in the context of traumatic experiences. The physical nature of the metaphors that bereaved parents employ when describing the intensity of their experiences and the fact that these metaphors sometimes spill over into their behaviour (e.g. the parent who literally removed her glasses) provide powerful insights into their physical and emotional state and as such are not simply ‘figures of speech’. These observations are also in line with previous research that has demonstrated that grief and emotional trauma may manifest as physical sensations[37].

Effective care and communication following child death takes account of the all-encompassing, frequently embodied nature of grief, and recognises that this may have an effect on parents’ abilities to make decisions and understand the meaning communicated in the care provided. Grief may manifest in behaviours that appear unusual or may go unnoticed (removing glasses), but are motivated by highly individualised metaphorical thinking patterns that represent the parents’ attempts to manage and come to terms with their experiences.

### Time

Following the death of a child, parents may perceive or experience time differently. Time might metaphorically expand or contract, or do both at the same time, and parents may feel that they are no longer part of the same ‘reality’ as those around them. While altered perceptions of time over the course of the grieving process have been attested to in narratives following child loss[38], our findings indicate that changed experiences of time in the immediate aftermath of a loss may also have important implications for care. It is therefore important for HCPs to recognise that parents’ emotional state may not be conducive to receiving and processing complex information, and they may need to be given the information more than once and/or in more than one format[32]. HCPs should demonstrate patience and sensitivity to parents’ difficulties in processing information.

There is a danger that care following the death of a child can become procedural, and as a result may not be empathetic to the needs of parents. For example, providing a memory box may constitute excellent bereavement care, but if this is given without clear explanation, or choice, may become a source of anxiety.. Following formal procedures without sensitivity to individual needs may result in greater disquiet.

This study has contributed to the growing literature on the experience of parental bereavement but its focus on the use of metaphor in parents’ accounts of child loss provides valuable insights. Key aspects of the experience have been identified that need to be considered when providing care for the bereaved. Our metaphorical analysis has afforded a focus on the on the language that was used to express content in qualitative interviews, providing more depth and nuance to interpretation. The number of participants was relatively small UK-based and self-selecting which could limit the transferability of our findings. Future studies could usefully explore metaphor with more participants with different demographic characteristics and in different settings.

## CONCLUSION

This study has shown how the analysis of metaphors used by parents when describing their experiences of bereavement can provide a deep and nuanced understanding of their experiences and how these experiences affect their evaluations of the communication they receive from HCPs. We have demonstrated that effective communication and strategies to ameloriate distress entails an understanding of the feelings and experiences of the bereaved and takes these into account. Conversely, ineffective communication is not grounded in this awareness.

Effective communication that takes account of parents’ experiences of bereavement may involve:

### Identity

- Demonstrating an awareness and acknowledgement of the parents’ identities as parents, and the child’s personhood
- Demonstrating respect for the child’s body and reassure parents that the child is being cared for even after death
- Facilitating opportunities for the parents to enact their roles as parents

### Emotional and Physical Responses

- Being aware that the all-encompassing nature of the grief may render parents unable to process and act upon information
- Recognising that parents may engage in individualised behaviours that may seem unusual or go unrecognised as enactments of grief.

### Time

- Recognising that parents may be experiencing time in altered ways, and that this may have an impact on their ability to process information.
- Explaining information in a clear but sensitive way, recognising that information may need to be given more than once or in different formats.
- Demonstrating patience, giving parents time and not rushing them through the various procedures or difficult decisions.

## Supporting information

COREQ checklist

## Data Availability

No data are available. The research involved interviews in which bereaved parents gave personal accounts of their experiences. Participants consented to their data being used for this project only. Therefore, we will not share the data for any other purpose.

## ACKNOWLEDGEMENTS

The authors thank the True Colours Trust for supporting this project.

## CONTRIBUTORS

All authors contributed equally to all stages of the process.

## PATIENT CONSENT FOR PUBLICATION

Obtained

## ETHICAL APPROVAL

The study had full ethical approval from University of Birmingham’s Ethical Review Committee (ERN_19-1582)

## PROVENANCE AND PEER REVIEW

Commissioned and funded by the True Colours Trust; grant number TCT1609; externally peer reviewed

