## supplemental table 1 for "Metaphors that shape parents’ perceptions of effective communication with healthcare practitioners following child death: a qualitative UK study"

**Supplementary material Table 1**

### Coding categories used in our analysis

01 Bad Communication
- a Dehumanisation of child
- b Lack of empathy
- c Use of jargon
- d Ineffective use of time
- e Wrong information provided
- f Bad advice
- g Failure to communicate key information

02 Good Communication
- a Recognition of child
- b Empathy
- c Clear language
- d Effective use of time
- e Correct information provided
- f Good advice
- g Successful communication of key information

03 Emotional Reactions
- Emotional reactions - negative
- Emotional reactions - neither positive or negative
- Emotional reactions - positive
- Unexpectedly powerful reactions - negative
- Unexpectedly powerful reactions - positive
- Wished for emotional reaction

04 Practical Reactions and Arrangements
- Practical reactions and arrangements - neither positive or negative
- Practical reactions and arrangements - positive
- Practical reactions and arrangements - negative
- Symbolic Behaviours
- Unexpectedly powerful reactions - negative
- Unexpectedly powerful reactions - positive
- Wished for practical reaction

05 People Involved
- GPs
- Hospital staff
- Doctors
- Nurses
- Others - hospital
- Receptionists
- Mortuary staff
- Paramedics

06 Continuing Bonds

07 Loss of Future

08 Defining Bereavement

09 Conceptions of time

10 Metaphors
11 Advice that parents would give
