## Supplementary material for "Metaphors that shape parents’ perceptions of effective communication with healthcare practitioners following child death: a qualitative UK study": COREQ checklist

### Parents' experiences of healthcare professional communication following the death of a child: A qualitative study in the UK

**Consolidated criteria for reporting qualitative studies (COREQ): 32-item checklist**

Developed from:
Tong A, Sainsbury P, Craig J. Consolidated criteria for reporting qualitative research (COREQ): a 32-item checklist for interviews and focus groups. *International Journal for Quality in Health Care*. 2007. Volume 19, Number 6: pp. 349 – 357

| No. Item | Guide questions/description | Reported on Page # |
|---|---|---|
| **Domain 1: Research team and reflexivity** | | |
| *Personal Characteristics* | | |
| 1. Inter viewer/facilitator | Which author/s conducted the interview or focus group?<br>All | 6 |
| 2. Credentials | What were the researcher's credentials?<br>ST - PhD in Applied Linguistics<br>JL - PhD in Applied Linguistics<br>AT - PhD in Nursing<br>JT - PhD in Nursing<br>EP - MA in Applied Linguistics | |
| 3. Occupation | What was their occupation at the time of the study?<br>ST - Lecturer in English<br>JL - Professor of Applied Linguistics<br>JT - Professor of Nursing<br>AT - Professor of Nursing<br>EP - Research assistant and postgrad student | |
| 4. Gender | Was the researcher male or female?<br>All female | |
| 5. Experience and training | What experience or training did the researcher have?<br>*All researchers trained in sensitive interviewing techniques*<br>*JL - experience of interviewing women who had experienced pregnancy loss* | |
| *Relationship with participants* | | |

| | | |
|---|---|---|
| 6. Relationship established | Was a relationship established prior to study commencement?<br>No | 5 |
| 7. Participant knowledge of the interviewer | What did the participants know about the researcher?<br>Place of work<br>Profession<br>Broad aims of the study | 5 |
| 8. Interviewer characteristics | What characteristics were reported about the interviewer/facilitator? | Name, gender, place of work |

| **Domain 2: study design** | | |
|---|---|---|
| *Theoretical framework* | | |
| 9. Methodological orientation and Theory | What methodological orientation was stated to underpin the study?<br>*Thematic analysis involving metaphor* | 6 |
| *Participant selection* | | |
| 10. Sampling | How were participants selected? e.g. purposive, convenience, consecutive, snowball.<br>*Purposive; advertisements were shared by relevant charities. Participants self-selected.* | 5 |
| 11. Method of approach | How were participants approached? e.g. face-to-face, telephone, mail, email-<br>*Recruitment through charities* | 5 |
| 12. Sample size | How many participants were in the study?<br>*24 across 21 interviews* | 7 |
| 13. Non-participation | How many people refused to participate or dropped out? Reasons?<br>*0 - participants self-selected* | 7 |
| *Setting* | | |
| 14. Setting of data collection | Where was the data collected?<br>*Face to face: In participants' homes or at the TCT HQ in London.*<br>*Online: Skype or Zoom* | 6 |
| 15. Presence of non-participants | Was anyone else present besides the participants and researchers?<br>*No* | 6 |
| 16. Description of sample | What are the important characteristics of the | |

| | | |
|---|---|---|
| | sample? e.g. demographic data, date<br>*All participants over 18 and having experienced child bereavement. Further demographic data not collected as not deemed necessary for study.* | |
| *Data collection* | | |
| 17. Interview guide | Were questions, prompts, guides provided by the authors? Was it pilot tested?<br>*Suggested open-ended interview prompts were provided; not piloted.* | 6 |
| 18. Repeat interviews | Were repeat interviews carried out? If yes, how many?<br>*No* | 6 |
| 19. Audio/visual recording | Did the research use audio or visual recording to collect the data?<br>*Audio recorded. Recordings on Skype/Zoom included video data which was destroyed after collection.* | 6 |
| 20. Field notes | Were field notes made during and/or after the interview or focus group?<br>*Yes* | 6 |
| 21. Duration | What was the duration of the interviews or focus group?<br>*Dependent on participant - range of 34 to 129 mins* | 6 |
| 22. Data saturation | Was data saturation discussed? No | n/a |
| 23. Transcripts returned | Were transcripts returned to participants for comment and/or correction?<br>Yes | 6 |
| **Domain 3: analysis and findings** | | |
| *Data analysis* | | |
| 24. Number of data coders | How many data coders coded the data?<br>*3* | |
| 25. Description of the coding tree | Did authors provide a description of the coding tree? yes | Online supplementary |
| 26. Derivation of themes | Were themes identified in advance or derived from the data?<br>*Derived from the data* | 7 |
| 27. Software | What software, if applicable, was used to manage the data? | 6 |

| | | |
|---|---|---|
| | *NVivo* | |
| 28. Participant checking | Did participants provide feedback on the findings?<br>*No* | |
| *Reporting* | | |
| 29. Quotations presented | Were participant quotations presented to illustrate the themes/findings? Was each quotation identified? e.g. participant number<br><br>*Quotes identified by participant code* | 7-10 |
| 30. Data and findings consistent | Was there consistency between the data presented and the findings?<br>*Yes* | |
| 31. Clarity of major themes | Were major themes clearly presented in the findings?<br>*Yes* | |
| 32. Clarity of minor themes | Is there a description of diverse cases or discussion of minor themes?<br>*No* | n/a |
